# Efficacy of Disease-Modifying Treatments in Transthyretin Amyloid Cardiomyopathy: A Systematic Review and Meta-Analysis of Five Phase III Randomized Controlled Trials

**DOI:** 10.64898/2026.10.10.26365208

**Authors:** Yi-Hsin Hung, Cheng-Hsuan Tsai, Ting-Wei Kao, Yen-Wen Wu, Wen-Chung Yu, Samuela Carigi, Brett W. Sperry, Stefano Perlini, Yen-Hung Lin

**Affiliations:** Division of Cardiology, Department of Internal Medicine, National Taiwan University Hospital and National Taiwan University College of Medicine, Taipei, Taiwan; Department of Internal Medicine, National Taiwan University Cancer Center, Taipei, Taiwan; Department of Internal Medicine, Far Eastern Memorial Hospital, New Taipei City, Taiwan; Division of Cardiology, Cardiovascular Medical Center, Far Eastern Memorial Hospital, New Taipei City, Taiwan; School of Medicine, National Yang Ming Chiao Tung University, Taipei, Taiwan; Department of Internal Medicine, College of Medicine, National Yang Ming Chiao Tung University, Taipei, Taiwan; Cardiovascular Center, Taipei Veterans General Hospital, Taipei, Taiwan; Cardiology Unit, Infermi Hospital, Rimini, Italy; Saint Luke’s Mid America Heart Institute, Kansas City, Missouri, USA; Department of Internal Medicine, Amyloid Research and Treatment Center, UOC Emergency Medicine, IRCCS Policlinico San Matteo Foundation, University of Pavia, Pavia, Italy

**Author notes:** These authors contributed equally to the study (Cheng-Hsuan Tsai, Yen-Hung Lin). **Corresponding author: Cheng-Hsuan Tsai**, Department of Internal Medicine, Division of Cardiology, National Taiwan University Hospital No. 7, Zhongshan S. Road, Zhongzheng District, Taipei 100, Taiwan., Email Address, **Yen-Hung Lin**, Department of Internal Medicine, Division of Cardiology, National Taiwan University Hospital, No. 7, Zhongshan S. Road, Zhongzheng District, Taipei 100, Taiwan.

**Keywords:** transthyretin amyloid cardiomyopathy, disease-modifying therapy, meta-analysis, randomized controlled trials

## Abstract

**Background:** Several disease-modifying agents for transthyretin amyloid cardiomyopathy (ATTR-CM) have been investigated, but differences in trial design and background stabilizer use complicate interpretation of their efficacy. This meta-analysis assessed their efficacy overall and by background stabilizer use.

**Methods:** PubMed, Embase, and the Cochrane Library were searched through September 2026 for phase III randomized, placebo-controlled trials of disease-modifying therapy in ATTR-CM. Primary endpoints were all-cause mortality, 6-minute walk distance (6MWD), Kansas City Cardiomyopathy Questionnaire-Overall Summary (KCCQ-OS) score, and N-terminal pro-B-type natriuretic peptide (NT-proBNP), pooled with a random-effects model. Treatment effects were compared between the monotherapy and baseline stabilizer strata using a mixed-effects model.

**Results:** Five trials (ATTR-ACT, APOLLO-B, ATTRibute-CM, HELIOS-B, CARDIO-TTRansform; N=3497) were included. Disease-modifying therapy reduced all-cause mortality (risk ratio, 0.76; 95% confidence interval [CI], 0.66-0.87; *I*^2^=0%) and improved 6MWD (pooled standardized mean difference [SMD], 0.43; 95% CI, 0.20-0.67), KCCQ-OS score (SMD, 0.37; 95% CI, 0.19-0.55), and NT-proBNP (SMD, - 0.53; 95% CI, −0.89 to −0.18). The mortality benefit was similar whether therapy was given alone or added to a background stabilizer (pooled risk ratio, 0.77 vs. 0.71; *P* for interaction = 0.59), but treatment effects on 6MWD (pooled SMD, 0.49 vs. 0.14), KCCQ-OS score (0.44 vs. 0.07), and NT-proBNP (−0.65 vs. −0.13) were smaller among patients receiving a background stabilizer (*P* for interaction = 0.012, <0.001, and 0.029, respectively).

**Conclusion:** Disease-modifying treatments reduced all-cause mortality and favored functional capacity, health status, and NT-proBNP outcomes in patients with ATTR-CM. Exploratory subgroup analyses showed smaller treatment effects on 6MWD, KCCQ-OS score, and NT-proBNP among patients receiving a background stabilizer, with no statistically significant difference in the mortality effect according to baseline stabilizer use.

**What Is New?:**

- This meta-analysis of all five phase III randomized controlled trials of disease-modifying therapy in transthyretin amyloid cardiomyopathy (3497 analyzed participants) found a lower risk of all-cause mortality (risk ratio, 0.76; 95% CI, 0.66-0.87) and more favorable 6-minute walk distance, Kansas City Cardiomyopathy Questionnaire–Overall Summary score, and N-terminal pro–B-type natriuretic peptide (NT-proBNP).
- In subgroup analyses, the mortality effect was similar with and without a background transthyretin stabilizer at randomization, whereas effects on functional capacity, health status, and NT-proBNP were smaller among patients receiving a background stabilizer.

**What Are the Clinical Implications?:**

- Among patients already receiving a transthyretin stabilizer, adding a second agent was associated with a mortality reduction similar to that seen without background therapy but with smaller incremental gains in functional and biomarker outcomes; because these findings rest on aggregate trial data and subgroup comparisons, they should be interpreted cautiously.

## INTRODUCTION

Transthyretin amyloid cardiomyopathy (ATTR-CM) is increasingly recognized as a significant cause of heart failure^1–6^. The pathogenesis of ATTR-CM begins with the pathological dissociation of transthyretin tetramers into monomers, which subsequently accumulate in the heart^7^. Its progressive and irreversible nature is associated with a poor long-term prognosis^8,9^. In recent years, various pharmacological treatments have been investigated, focusing on different therapeutic targets^10–14^. These targets include reducing transthyretin production, preventing the dissociation of the transthyretin tetramer, and depleting established transthyretin amyloid deposits^15–21^.

Transthyretin stabilizers, including tafamidis and acoramidis, prevent the dissociation of transthyretin tetramers into monomers and thereby mitigate the progression of ATTR-CM^15,17^. These medications were studied in the Tafamidis in Transthyretin Cardiomyopathy Clinical Trial (ATTR-ACT) and in the Efficacy and Safety of Acoramidis in Subjects With Transthyretin Amyloid Cardiomyopathy (ATTRibute-CM) trial^15,17^. Gene-silencing agents act instead by reducing hepatic transthyretin production^16,18,19^. Patisiran and vutrisiran are small interfering RNAs and were studied in the Patisiran Treatment in Patients with Transthyretin Cardiac Amyloidosis (APOLLO-B) trial and in the Patients with Transthyretin Amyloidosis with Cardiomyopathy (HELIOS-B) trial, respectively^16,18^. Eplontersen is a ligand-conjugated antisense oligonucleotide and was studied in the Eplontersen for Transthyretin Amyloid Cardiomyopathy (CARDIO-TTRansform) trial, which was reported most recently and is the largest trial in this field to date^19^.

Although most of these agents met their designated primary endpoints, the trials differed substantially in design, and their results are not directly comparable. The primary endpoints were not the same: the 6-minute walk distance (6MWD) at 12 months in APOLLO-B, hierarchical composites of death and cardiovascular hospitalization in ATTR-ACT and ATTRibute-CM, and composites of death and recurrent cardiovascular events in HELIOS-B and CARDIO-TTRansform^15–19^. Follow-up ranged from 12 to approximately 36 months^15–19^. The trials also differed in a respect that has become central to contemporary practice. In ATTR-ACT and ATTRibute-CM, no participants were receiving a background stabilizer at randomization; these trials therefore evaluated the initiation of a disease-modifying agent versus placebo^15,17^. By contrast, approximately 25%, 40% and 57% of participants in APOLLO-B, HELIOS-B and CARDIO-TTRansform, respectively, were already receiving a stabilizer at randomization, allowing assessment of the incremental effects of gene-silencing therapy in these patients^16,18,19^.

Several meta-analyses of disease-modifying therapy in ATTR-CM have been published^22,23^. However, none has included all five randomized controlled trials. We therefore conducted this meta-analysis of all five phase 3 randomized, placebo-controlled trials to evaluate the overall efficacy of disease-modifying therapy in ATTR-CM and explore differences in treatment effects according to background transthyretin stabilizer use at baseline.

## METHODS

### Study registration

This systematic review and meta-analysis were registered with the International Platform of Registered Systematic Review and Meta-Analysis Protocols (INPLASY; registration number INPLASY202690037) and was conducted in adherence to the PRISMA (Preferred Reporting Items for Systematic Reviews and Meta-Analyses) guidelines. The Cochrane risk-of-bias tool for randomized trials was used to assess risk of bias.

### Search strategy

PubMed, Embase, and the Cochrane Library were searched for eligible studies from inception to September 30^th^, 2026, without language restriction. Only phase 3 randomized controlled trials in human subjects investigating disease-modifying therapy for ATTR-CM were eligible; trials investigating target-organ involvement other than cardiomyopathy were excluded. Medical subject headings and keywords included (“Transthyretin Amyloidosis” OR “Transthyretin Amyloid Cardiomyopathy”) AND (“cardiomyopathy”) AND (“Trial”) AND (“Efficacy” OR “Treatment”) AND (“Randomized”) AND (“Trial”) NOT (“Review”), as shown in **Table S1**. Literature screening followed the PRISMA algorithm. Following the original search (through October 5, 2024), the search was re-run to incorporate CARDIO-TTRansform, the most recently reported trial in this field.

### Clinical endpoints

The endpoints of interest were all-cause mortality, the 6MWD, the Kansas City Cardiomyopathy Questionnaire-Overall Summary (KCCQ-OS) score, and N-terminal pro-B-type natriuretic peptide (NT-proBNP), each assessed at the longest available follow-up in each trial. Analyses were performed in three prespecified cohorts: (1) the oveall analysis population; (2) the monotherapy stratum, defined by the absence of a background transthyretin stabilizer at randomization; and (3) the combination stratum, defined by ongoing transthyretin stabilizer use at randomization. The therapies evaluated were transthyretin stabilizers in ATTR-ACT and ATTRibute-CM and gene silencers in APOLLO-B, HELIOS-B, and CARDIO-TTRansform.

### Data extraction

Titles, abstracts, and full-text articles were reviewed independently by two authors (Y.H. Hung and C.H. Tsai); disagreements at any stage were resolved by consensus. Extracted data included study duration, medication type, number of participants, age, sex, ATTR mutation status (wild-type versus variant), and New York Heart Association (NYHA) functional class. For all-cause mortality, the number of events and the number of participants in each randomized arm were extracted. For continuous outcomes (6MWD, KCCQ-OS score, and NT-proBNP), extracted data included means with standard deviations, medians with interquartile ranges, medians with 95% CIs, least-squares means with standard errors or 95% CIs, and ratios of adjusted geometric means with 95% CIs, as reported by each trial. Priority was given to trials reporting both pretreatment and post-treatment values; where only the change from baseline was reported, that value was extracted instead. Data were extracted separately for the overall population and, where reported, for the monotherapy and combination strata.

### Statistical analysis

A random-effects model, using the DerSimonian and Laird method, was applied to pool the results of comparing treatment arm to placebo arm from individual studies. All-cause mortality was pooled as a risk ratio on the log scale. Because continuous outcomes were reported on heterogeneous metrics across trials, values for 6MWD, KCCQ-OS, and NT-proBNP were transformed into standardized mean differences (SMDs) before pooling. Heterogeneity among trials was assessed with the *I*^2^ statistics and Cochran’s Q test. Differences in treatment effect between the monotherapy and combination strata were assessed using a mixed-effects model, with treatment stratum specified as a fixed effect. Publication bias was assessed qualitatively with funnel plots and quantitatively with the Egger test for each outcome. A two-sided *P* value < 0.05 was considered statistically significant. Analyses were performed using Comprehensive Meta-Analysis, Version 3 (Englewood, NJ 2014).

## RESULTS

### Included Studies and Risk of Bias Assessment

A total of 377 records were identified during the initial literature search. After removal of duplicate records, 199 records were screened by title and abstract, of which 138 were excluded. The remaining 61 full-text articles were assessed for eligibility. Ultimately, five phase 3 randomized controlled trials (ATTR-ACT, APOLLO-B, ATTRibute-CM, HELIOS-B and CARDIO-TTRansform) regarding the efficacy of disease-modifying treatments were pooled for meta-analysis. All studies were performed in a double-blinded, placebo-controlled manner. The algorithm of study inclusion is illustrated in **Figure 1**.

**Figure 1.**
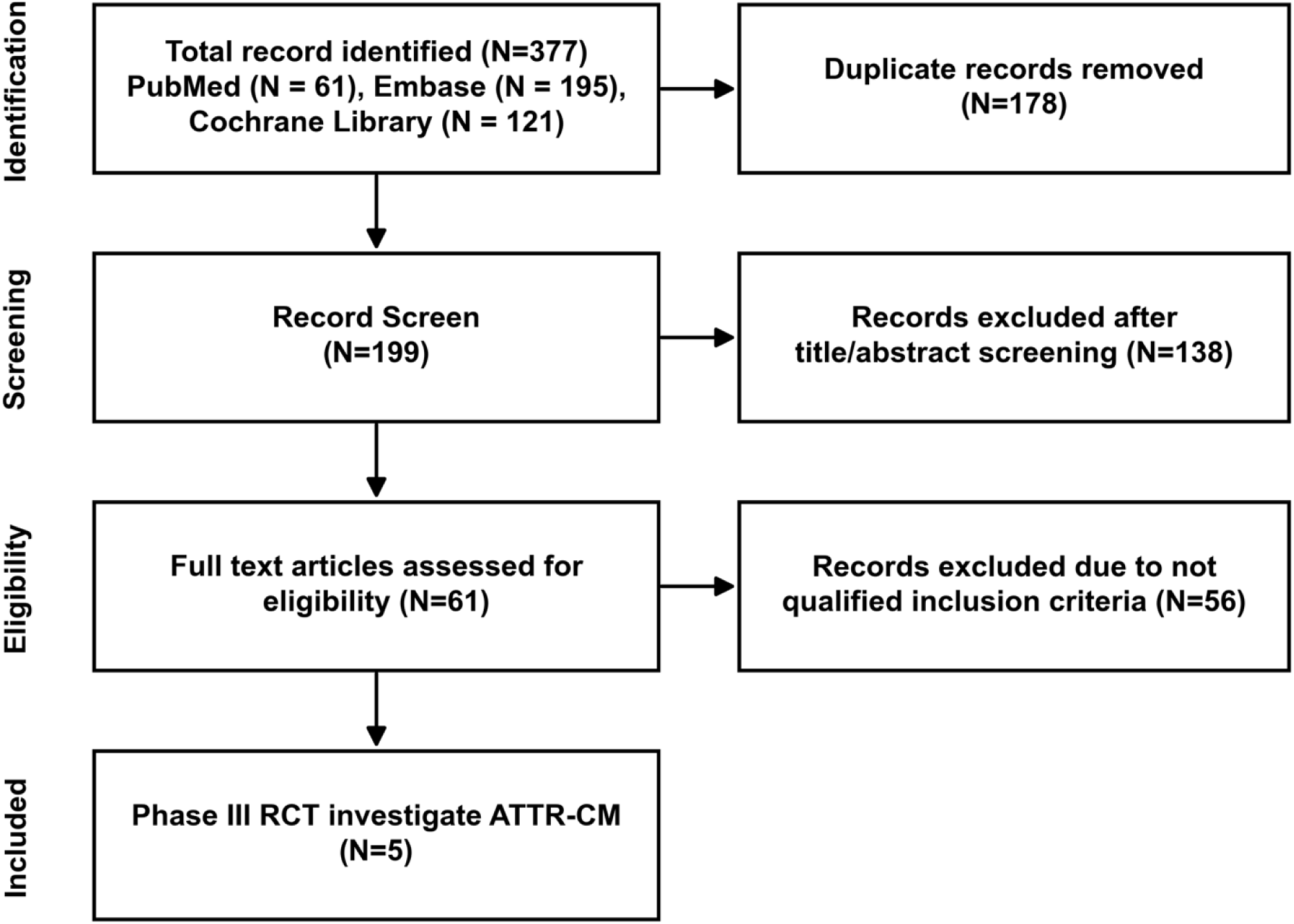
Flow diagram of study selection. PRISMA; Preferred Reporting Items for Systematic Reviews and Meta-Analyses; RCT, randomized controlled trial.

Overall, the risk of bias for these five randomized controlled trials was low (**Table S2**). The assessment of publication bias was provided in the Supplemental Material; no substantial evidence of small-study bias was observed (**Figure S1**).

### Baseline Characteristics

Overall, 3497 participants were included in the analyses from five trials evaluating tafamidis (ATTR-ACT; n = 441), patisiran (APOLLO-B; n = 359), acoramidis (ATTRibute-CM; n = 611), vutrisiran (HELIOS-B; n = 654) and eplontersen (CARDIO-TTRansform; n = 1432); 1895 participants were assigned to an active disease-modifying agent and 1602 to placebo. Follow-up was 30 months in ATTR-ACT and ATTRibute-CM, 12 months in APOLLO-B, 33 to 36 months in HELIOS-B and 140 weeks in CARDIO-TTRansform. Baseline characteristics of the included trials are shown in **Table 1**.

**Table 1.** Key Study Design Elements, Baseline Characteristics, and Background Medical Therapy of the Five Included Trials.

| Characteristic | ATTR-ACT<br>(n = 441) | APOLLO-B<br>(n = 359) | ATTRibute-CM<br>(n = 611) | HELIOS-B<br>(n = 654) | CARDIO-TTTransform<br>(n = 1432) |
| --- | --- | --- | --- | --- | --- |
| **Study design** |  |  |  |  |  |
| Comparison | Tafamidis (80 or 20 mg) vs placebo | Patisiran vs placebo | Acoramidis vs placebo | Vutrisiran vs placebo | Eplontersen vs placebo |
| Enrollment period | Dec 2013-Aug 2015 | Oct 2019-May 2021 | Apr 2019-Oct 2020 | Dec 2019-Aug 2021 | Mar 2020-Jul 2023 |
| **Key eligibility criteria** |  |  |  |  |  |
| Age, y | 18-90 | 18-85 | 18-90 | 18-85 | 18-90 |
| NYHA class/UK NAC stage | Excluded if NYHA class IV; NAC staging not used | Excluded if NYHA class IV, or class III with NAC stage 3 | Excluded if eGFR <15 mL/min/1.73 m<sup>2</sup>; randomization stratified by genotype, NT-proBNP, and eGFR (cut-offs corresponding to NAC staging) | Excluded if NYHA class IV, or class III with NAC stage 3 | Excluded if NYHA class IV |
| NT-proBNP, ng/L | ≥600; no upper limit | >300 (>600 in AF); <8500 | ≥300; excluded if ≥8500 | >300 (>600 in AF); <8500 | ≥600 (≥1200 in AF); no upper limit |
| 6-min walk distance, m | >100 | ≥150 | ≥150 on 2 tests 24 h-3 weeks apart | ≥150 | ≥100 |
| **End point** |  |  |  |  |  |
| Primary efficacy end point | Hierarchical Finkelstein-Schoenfeld: all-cause mortality, then frequency of CV-related hospitalization | Change from baseline in 6-min walk distance at 12 months | 4-step hierarchical Finkelstein-Schoenfeld: death, CV-related hospitalization, NT-proBNP, 6-min walk distance | Composite of all-cause death and recurrent CV events | Composite of CV death and recurrent CV events up to week 140 |
| Study duration | 30 months | 12 months | 30 months | 33-36 months | 140 weeks |
| **Patient demographics** |  |  |  |  |  |
| Age, y | 74.3 (7.0) | 76 (41-85) | 77.3 (6.6) | 77 (45-85) | 76.4 (7.1) |
| Men, No. (%) | 398 (90.2) | 321 (89.4) | 570 (90.2) | 605 (92.5) | 1297 (90.6) |
| TTR genotype, No. (%) |  |  |  |  |  |
| Wild-type ATTR | 335 (76.0) | 288 (80.2) | 571 (90.3) | 578 (88.4) | 1221 (85.3) |
| Variant ATTR | 106 (24.0) | 71 (19.8) | 61 (9.7) | 76 (11.6) | 211 (14.7) |
| **Disease severity** |  |  |  |  |  |
| NYHA class, No. (%) |  |  |  |  |  |
| I | 37 (8.4) | 25 (7.0) | 68 (10.8) | 84 (12.8) | 192 (13.4) |
| II | 263 (59.6) | 306 (85.2) | 455 (72.0) | 508 (77.7) | 997 (69.6) |
| III | 141 (32.0) | 28 (7.8) | 109 (17.2) | 62 (9.5) | 243 (17.0) |
| 6-min walk distance, m | NR | 358.0 (295.0-420.0) /<br>367.7 (300.0-444.3)* | NR | 374.6 (100.0) | 352.2 (96.2) |
| KCCQ-OS score | NR | 70.0 (20.9) | NR | 72.6 (19.7) | 72.6 (19.9) |
| NT-proBNP, median<br>(IQR), ng/L, active /<br>placebo | 2996 (1752-4862) / 3161 (1864-4825) | 2008 (1135-2921) /<br>1813 (952-3079) | 2326 (1332-4019) / 2306<br>(1128-3754) | 2021 (1138-3312) /<br>1801 (1042-3082) | 2068 (1323-3563) / 1965<br>(1284-3422) |
| **Background medical**<br>**therapy, No. (%)** |  |  |  |  |  |
| TTR stabilizer at baseline | 0 | 91 (25.3) | 0 | 259 (39.6) | 819 (57.2) |
| SGLT2 inhibitor | NR | 15 (4.2) | NR | 21 (3.2) | 265 (18.5) |
| Mineralocorticoid<br>receptor antagonist | NR | 166 (46.2) | NR | 230 (35.2) | 618 (43.2) |
| β-Blocker | NR | 150 (41.8) | NR | 288 (44.0) | 614 (42.9) |
| ACEi/ARB/ARNi | NR | 153 (42.6) | NR | 302 (46.2) | 591 (41.3) |
| Loop diuretic | NR | 332 (92.5) | NR | 507 (77.5) | 1243 (86.8) |

ACEi, angiotensin-converting enzyme inhibitor; AF, atrial fibrillation; ARB, angiotensin receptor blocker; ARNi, angiotensin receptor–neprilysin inhibitor; ATTR, transthyretin amyloidosis; CV, cardiovascular; eGFR, estimated glomerular filtration rate; IQR, interquartile range; KCCQ-OS, Kansas City Cardiomyopathy Questionnaire–Overall Summary; NAC, National Amyloidosis Centre; NR, not reported; NT-proBNP, N-terminal pro–B-type natriuretic peptide; NYHA, New York Heart Association; SGLT2, sodium-glucose cotransporter 2; TTR, transthyretin.
\*For APOLLO-B, the 6-min walk distance is shown as the median (IQR) in the active and placebo groups.

Across the five trials, the mean age was 74.3 years in ATTR-ACT and 77.3 years in ATTRibute-CM and the median age was 76 to 77 years in the other three trials; 89.4% to 92.5% of participants were men, and 76.0% to 90.3% had wild-type ATTR-CM. NYHA class III symptoms were present in 32.0% of ATTR-ACT participants, compared with 7.8% in APOLLO-B, 17.2% in ATTRibute-CM, 9.5% in HELIOS-B and 17.0% in CARDIO-TTRansform. The median NT-proBNP concentration was approximately 3000 ng/L in ATTR-ACT, compared with 1801 to 2326 ng/L in the four later trials.

Baseline use of concomitant TTR stabilizer therapy differed across the trials. No participant was receiving background disease-modifying treatment at randomization in ATTR-ACT or ATTRibute-CM, whereas 25.3% of participants in APOLLO-B, 39.6% in HELIOS-B and 57.2% in CARDIO-TTRansform were receiving a stabilizer at randomization. Overall, 2328 participants received disease-modifying monotherapy and 1169 received combination therapy on a background stabilizer.

### Overall Treatment Effects

All-cause mortality occurred in 323 of 1895 participants (17.0%) assigned to active therapy and in 337 of 1602 (21.0%) assigned to placebo. Disease-modifying therapy reduced all-cause mortality (pooled risk ratio, 0.76; 95% CI, 0.66-0.87), without evidence of heterogeneity between trials (*I*^2^ = 0.0%, *P* = 0.57) (**Figure 2A**).

**Figure 2.**
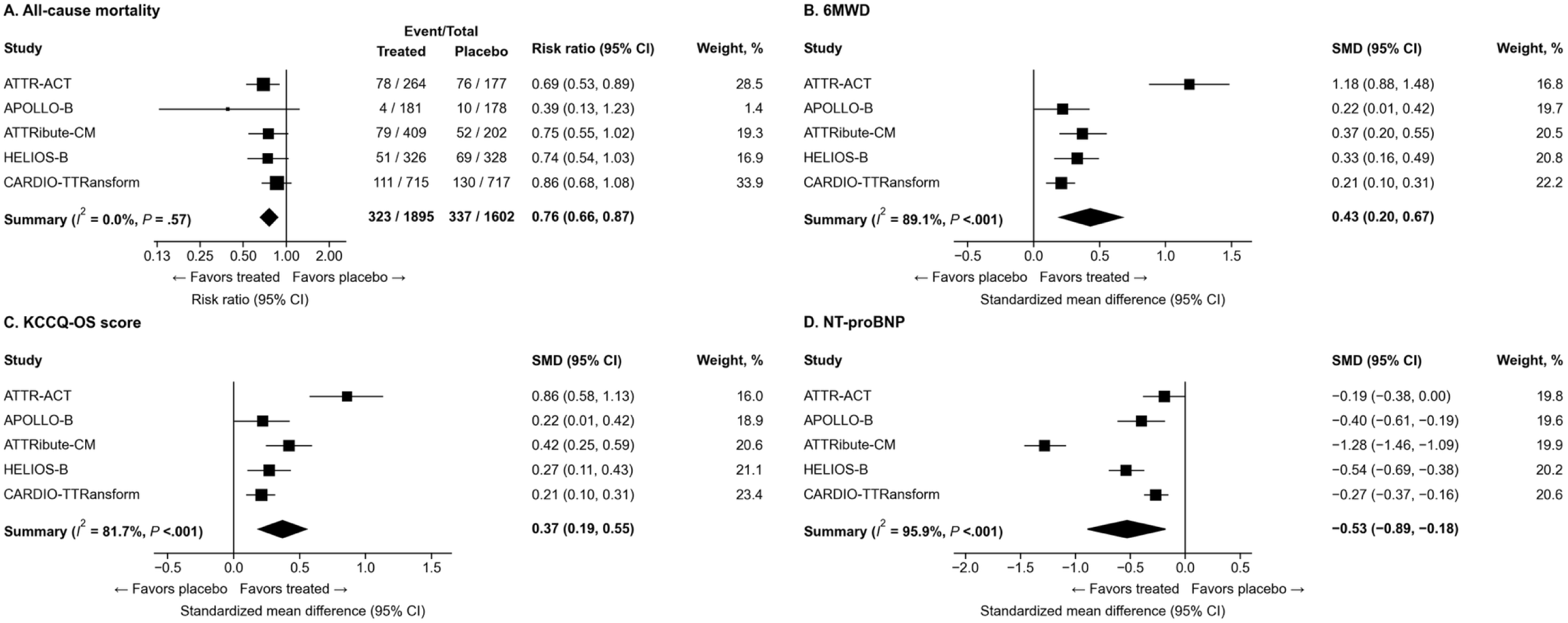
Effect of disease-modifying therapy versus placebo in the overall population. Forest plots show (A) all-cause mortality, expressed as risk ratio, and (B) 6-minute walk distance (6MWD), (C) Kansas City Cardiomyopathy Questionnaire-Overall Summary (KCCQ-OS) score, and (D) N-terminal pro-B-type natriuretic peptide (NT-proBNP), expressed as standardized mean difference (SMD). Each trial contributes its longest available follow-up.

Compared with placebo, disease-modifying therapy preserved functional capacity, as assessed by the 6MWD (pooled SMD, 0.43; 95% CI, 0.20-0.67) and the KCCQ-OS score (0.37; 95% CI, 0.19-0.55), and attenuated the rise in NT-proBNP (−0.53; 95% CI, −0.89 to −0.18) (**Figure 2B-D**). Between-trial heterogeneity was present for all three outcomes (*I*^2^ = 89.1%, 81.7% and 95.9%, respectively; *P* < 0.001 for each). Treatment effect estimates varied across trials. For 6MWD and KCCQ-OS score, the largest SMDs were observed in ATTR-ACT: 1.18 (95% CI, 0.88-1.48) and 0.86 (95% CI, 0.58-1.13), respectively. Corresponding SMDs in the other four trials ranged from 0.21 to 0.37 for 6MWD and from 0.21 to 0.42 for KCCQ-OS. For NT-proBNP, SMDs ranged from −1.28 in ATTRibute-CM to −0.19 in ATTR-ACT.

### Monotherapy versus Combination Therapy

Death from any cause occurred in 263 of 1309 participants (20.1%) assigned to active therapy and in 252 of 1019 (24.7%) assigned to placebo in the monotherapy stratum, and in 60 of 586 (10.2%) and 85 of 583 (14.6%), respectively, in the combination stratum. The effect on all-cause mortality did not differ according to background stabilizer use (pooled risk ratio, 0.77 [95% CI, 0.67-0.90] with monotherapy vs. 0.71 [95% CI, 0.52-0.96] with combination therapy; *P* for interaction = 0.59), with *I*^2^ = 0% in both strata (**Figure 3A**).

**Figure 3.**
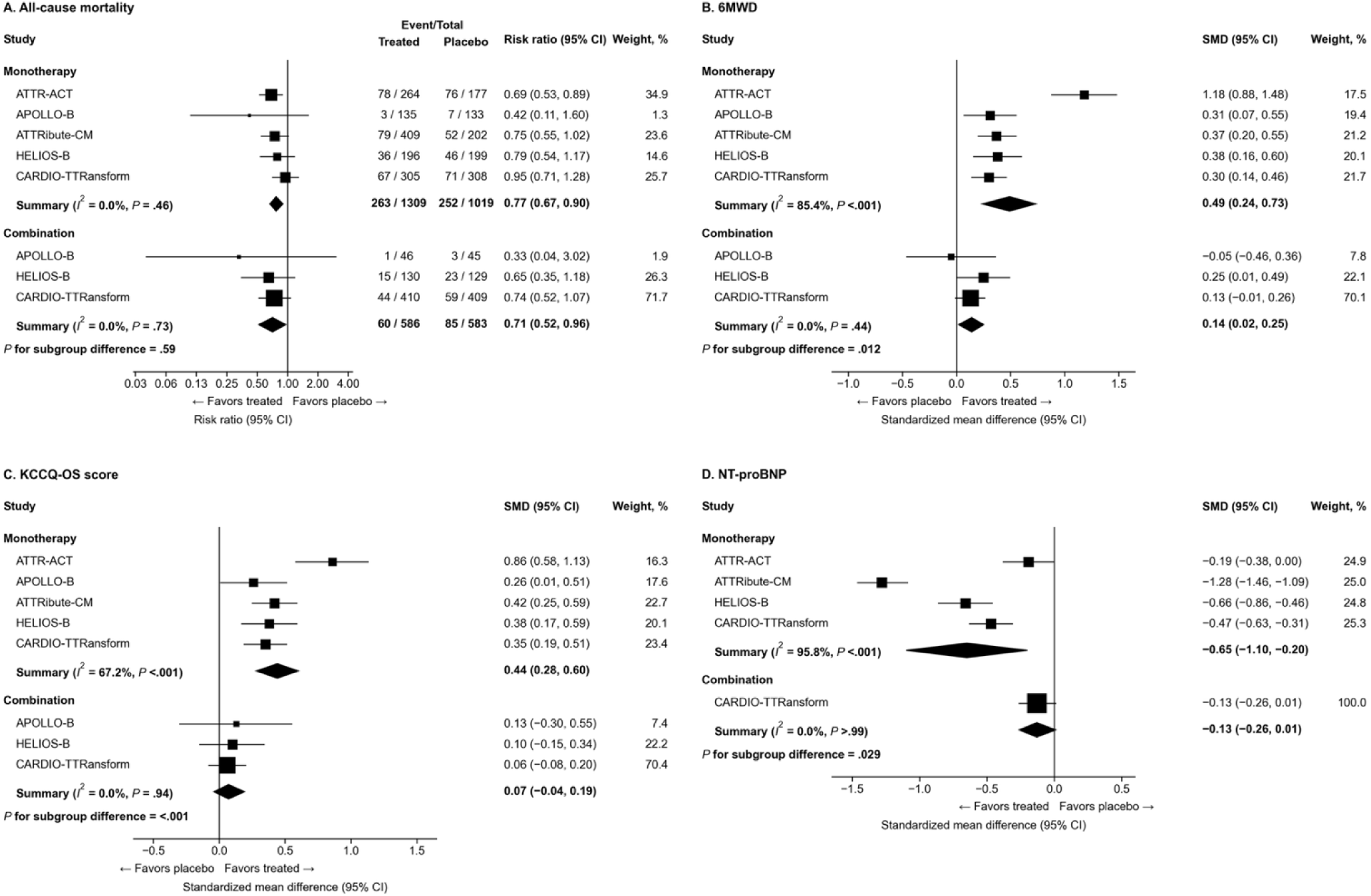
Effect of disease-modifying therapy versus placebo according to background transthyretin stabilizer use at randomization. Forest plots show (A) all-cause mortality (risk ratio), and (B) 6-minute walk distance (6MWD), (C) KCCQ-OS score, and (D) NT-proBNP (standardized mean difference [SMD]) in the monotherapy stratum (no background stabilizer) and the combination stratum (background stabilizer).

For the three remaining endpoints, treatment effects were significantly smaller in the combination stratum. The pooled SMD for the 6MWD was 0.49 (95% CI, 0.24-0.73) with monotherapy and 0.14 (95% CI, 0.02-0.25) with combination therapy (*P* for interaction = 0.012) (**Figure 3B**). For the KCCQ-OS score, the pooled SMD was 0.44 (95% CI, 0.28-0.60) and 0.07 (95% CI, −0.04 to 0.19) (*P* for interaction < 0.001) (**Figure 3C**). For NT-proBNP, the pooled SMD was −0.65 (95% CI, −1.10 to −0.20) and −0.13 (95% CI, −0.26 to 0.01) (*P* for interaction = 0.029) (**Figure 3D**). The combination stratum for NT-proBNP comprised CARDIO-TTRansform alone, as the remaining trials either had no combination population or did not report NT-proBNP according to baseline stabilizer use. For these three endpoints, between-trial heterogeneity was present in the monotherapy stratum (*I*^2^ > 50% for each) but not in the combination stratum (*I*^2^ = 0.0% for all three).

## DISCUSSION

This meta-analysis is the first to include all five phase 3, placebo-controlled trials of disease-modifying therapy in ATTR-CM. Across 3497 analyzed participants, disease-modifying therapy reduced all-cause mortality by 24%, improved the 6MWDand the KCCQ-OS score, and attenuated the rise in NT-proBNP. These effects were not uniform: among patients already taking a transthyretin stabilizer at randomization, adding a second agent preserved the reduction in mortality but was associated with smaller incremental gains in 6MWD, KCCQ-OS score and NT-proBNP than were seen in patients receiving no background treatment (**Central Illustration**).

In both strata the randomized comparison is active therapy versus placebo; what differs is the background therapy received by both arms. In the monotherapy stratum, neither arm was taking a stabilizer, so the estimate reflects the effect of a disease-modifying agent given alone. In the combination stratum, both arms were taking a stabilizer. Therefore, the comparison estimates the incremental effect of adding a gene-silencing agent to established stabilizer therapy, rather than the effect of combination therapy versus monotherapy. On that basis, there was no detected difference in the relative reduction in all-cause mortality between the two strata (risk ratio, 0.71 with combination therapy vs 0.77 with monotherapy), whereas treatment effects on 6MWD, KCCQ-OS score, and NT-proBNP were significantly smaller in the combination stratum. One possible explanation is that prior stabilizer treatment may limit the additional benefit achievable with gene silencing on functional capacity, health status, and NT-proBNP. However, differences in baseline disease severity, disease duration, and trial populations may also contribute. In particular, ATTR-ACT and ATTRibute-CM contributed exclusively to the stratum without background stabilizer therapy, meaning that comparisons between strata also reflect differences in trial composition and drug mechanism^15,17^. The aggregate data cannot distinguish among these explanations. These findings therefore do not establish that combination therapy is less effective than monotherapy or identify the preferred initial treatment strategy. Individual-participant data analyses and randomized comparisons of treatment strategies are needed to clarify which patients may benefit from adding a gene silencer to stabilizer therapy^24–27^.

These findings should also be considered in the context of the evolving ATTR-CM trial landscape. ATTR-ACT recruited between 2013 and 2015 and enrolled the most advanced disease of the five: NYHA class III symptoms were nearly twice as common as in any later trial, and the median NT-proBNP concentration was approximately 30-60% higher (**Table 1**)^15^. Consistent with these differences, among control participants not receiving a stabilizer at baseline, mortality over the trial follow-up was lower in the more contemporary trials (ATTRibute-CM, 25.7% at 30 months; HELIOS-B, 23.1% over the double-blind period; and CARDIO-TTRansform, 23.1% at week 140) than in ATTR-ACT (42.4% at 30 months) (**Figure 4**)^15,17,19^. Background heart failure therapy also changed across the enrollment periods, most notably for sodium-glucose cotransporter 2 (SGLT2) inhibitors, taken at baseline by 3.2% of HELIOS-B participants and 18.5% of those in CARDIO-TTRansform^18,19^. This may be a clinically relevant difference, as SGLT2 inhibitor use has itself been associated with better outcomes in patients with ATTR-CM^28–30^. These temporal and clinical differences may contribute to between-trial heterogeneity and should be considered when comparing treatment-effect estimates across studies.

**Figure 4.**
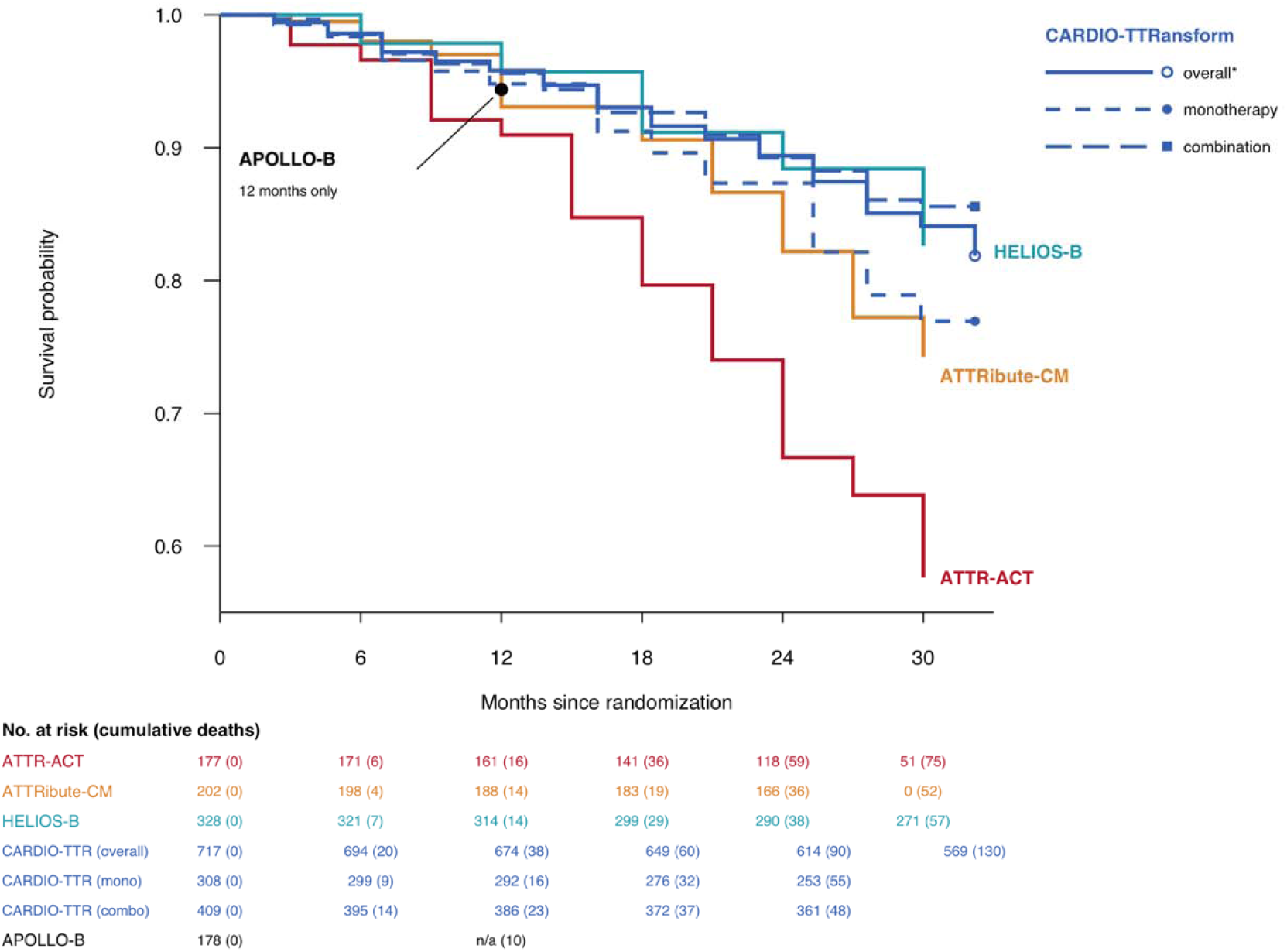
All-cause mortality in the placebo groups of the included trials. Curves show survival probability derived from the number-at-risk and cumulative-event tables in the original publications. *The CARDIO-TTRansform overall curve was estimated from the published decline in number at risk, calibrated to the reported 30-month death count; the monotherapy and combination curves were derived from published number-at-risk tables.

Our findings are consistent with and extend those of previous meta-analyses^22–24^. Autherith et al. pooled the four trials available before CARDIO-TTRansform and demonstrated an overall benefit of disease-modifying therapy, although treatment effects were not stratified by background stabilizer use^22^. Gillmore et al. subsequently examined background stabilizer use in HELIOS-B and CARDIO-TTRansform and similarly found smaller treatment effects on 6MWD and KCCQ-OS score among patients receiving a background stabilizer, without significant heterogeneity in the effect on all-cause mortality^23^. Our analysis extends this evidence by including all five phase 3 trials, encompassing 3497 participants across both stabilizer and gene-silencing therapies and providing a more comprehensive assessment of disease-modifying therapy in the evolving ATTR-CM treatment landscape.

### Limitations

This study has several limitations. First, the analysis relied on published aggregate data rather than individual-participant data, limiting our ability to account for differences in patient characteristics and assess treatment-effect modification. The trials also differed in follow-up duration, outcome assessment, and analytical methods, which may affect the comparability of the pooled estimates. Second, participants were classified according to stabilizer use at randomization, but some patients initiated stabilizer therapy during follow-up. Thus, baseline treatment status did not fully capture treatment exposure throughout the trials, which may complicate interpretation of the subgroup findings. Third, differences in baseline demographics across the trials also influenced the observed differences in clinical outcomes. Despite the potential for inaccuracies, the pooled estimate from this meta-analysis remains valuable for synthesizing the current evidence. Fourth, substantial heterogeneity was observed for the continuous outcomes, warranting caution when interpreting their pooled effect sizes. Finally, subgroup data were limited for some outcomes, particularly NT-proBNP, for which only one trial contributed data among patients receiving a background stabilizer. The small number of included trials also precluded a reliable assessment of publication bias.

## CONCLUSION

In this meta-analysis of all five phase 3 placebo-controlled trials in ATTR-CM, disease-modifying therapy reduced all-cause mortality, improved the 6MWD and the KCCQ-OS score, and attenuated the rise in NT-proBNP. Among patients already receiving a transthyretin stabilizer, adding a second agent preserved the mortality benefit but was associated with smaller treatment effects on the other three endpoints. Further studies are needed to clarify the incremental benefit of adding gene-silencing therapy to stabilizer treatment and identify patients most likely to benefit.

## Data Availability

All data analyzed in this study were obtained from publicly available published randomized controlled trials. The data supporting the findings of this study are available in the original publications and their supplementary materials.

## Acknowledgments

None

**Supplemental Material**

**Tables S1–S2**

**Figure S1**

## Non-standard Abbreviations and Acronyms

6MWD: 6-minute walk distance
ATTR-CM: transthyretin amyloid cardiomyopathy
KCCQ-OS: Kansas City Cardiomyopathy Questionnaire-Overall Summary
NT-proBNP: N-terminal pro-B-type natriuretic peptide
SGLT2: sodium-glucose cotransporter 2
SMD: standardized mean difference
TTR: transthyretin

---

**Central Illustration.**
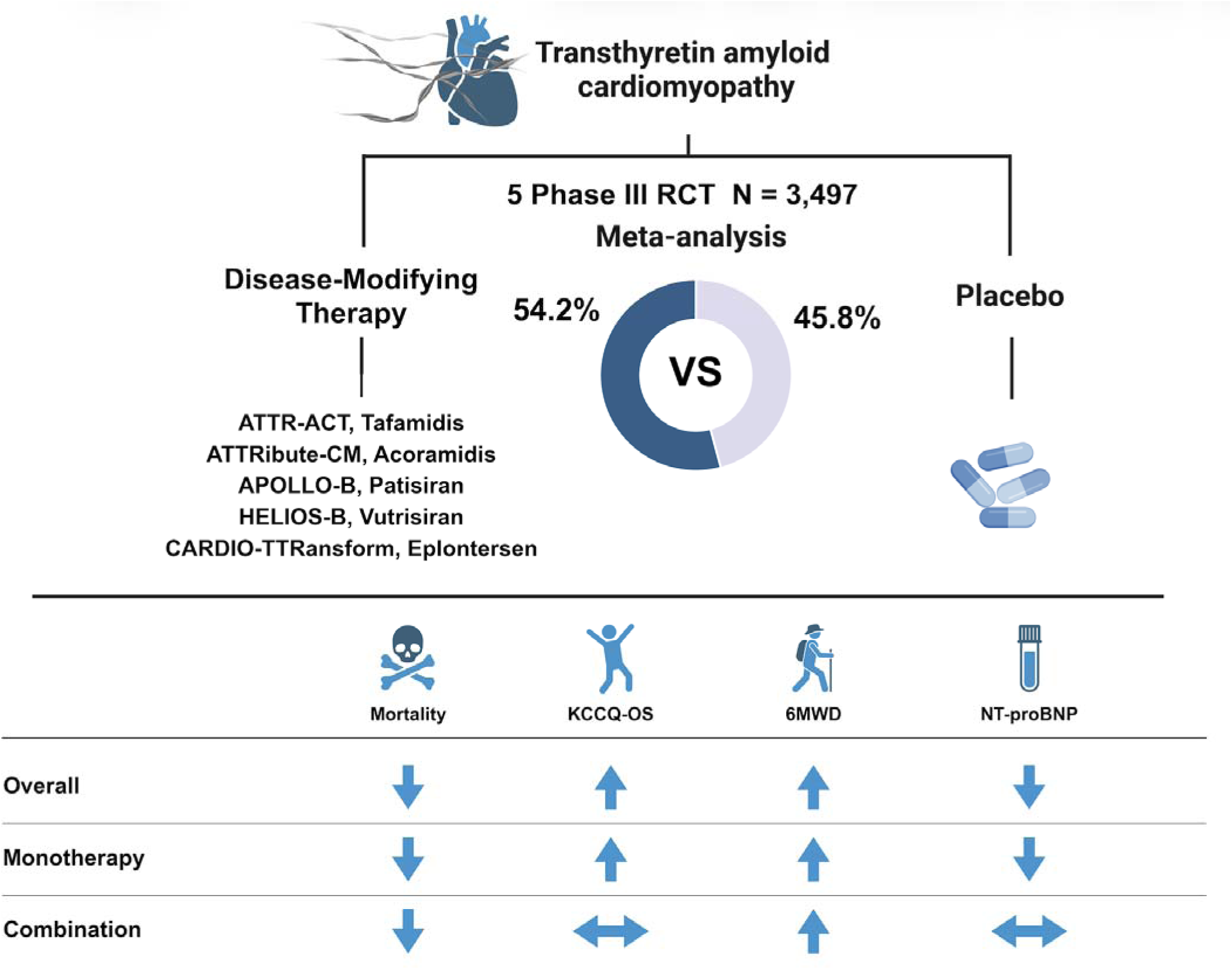
Disease-modifying therapy in transthyretin amyloid cardiomyopathy: summary of a meta-analysis of five phase III randomized controlled trials. Strata are defined by transthyretin stabilizer use at randomization. In the monotherapy (no-baseline-stabilizer) stratum, the comparison is a disease-modifying agent versus placebo. In the combination (baseline-stabilizer) stratum, all patients were already receiving a stabilizer, so the comparison estimates the incremental effect of adding a gene-silencing agent to a stabilizer versus adding placebo; only APOLLO-B, HELIOS-B and CARDIO-TTRansform contributed to this stratum. ↔ indicates that the 95% confidence interval includes no difference. KCCQ-OS, Kansas City Cardiomyopathy Questionnaire-Overall Summary; NT-proBNP, N-terminal pro–B-type natriuretic peptide; 6MWD, 6-minute walk distance.

