## Supplemental data for "Efficacy of Disease-Modifying Treatments in Transthyretin Amyloid Cardiomyopathy: A Systematic Review and Meta-Analysis of Five Phase III Randomized Controlled Trials"

**Supplemental Table 1.** Search Strategy and Keywords Used for the Systematic Literature Search

| Database | Search syntax |
| --- | --- |
| PubMed | ("Transthyretin Amyloidosis" or "Transthyretin Amyloid Cardiomyopathy") AND ("cardiomyopathy") AND ("Trial") AND ("Efficacy" OR "Treatment") AND ("Randomized") AND ("Trial") NOT (“Review”) |
| EMBASE | ("Transthyretin Amyloidosis" or "Transthyretin Amyloid Cardiomyopathy") AND ("cardiomyopathy") AND ("Trial") AND ("Efficacy" OR "Treatment") AND ("Randomized") AND ("Trial") NOT (“Review”) |
| Cochrane’s library | ("Transthyretin Amyloidosis" or "Transthyretin Amyloid Cardiomyopathy") AND ("cardiomyopathy") AND ("Trial") AND ("Efficacy" OR "Treatment") AND ("Randomized") AND ("Trial") NOT (“Review”) |

**Supplemental table 2.** Randomized controlled trials were appraised by Cochrane Risk of Bias

| Studies | Sequence generation | Allocation concealment | Blinding | Incomplete results | Selective reporting | Other bias |
| --- | --- | --- | --- | --- | --- | --- |
| Maurer, 2018 | Low | Low | Low | Low | Low | Low |
| Maurer, 2023 | Low | Low | Low | Low | Low | Low |
| Gillmore, 2024 | Low | Low | Low | Low | Low | Low |
| Fontana, 2024 | Low | Low | Low | Low | Low | Low |
| Fontana, 2026 | Low | Low | Low | Low | Low | Low |

**Supplemental Figure 1**. Funnel plots for assessment of publication bias. Panels show (A) all-cause mortality (log risk ratio), and (B) 6-minute walk distance (6MWD), (C) KCCQ-OS score, and (D) NT-proBNP (standardized mean difference) plotted against standard error.

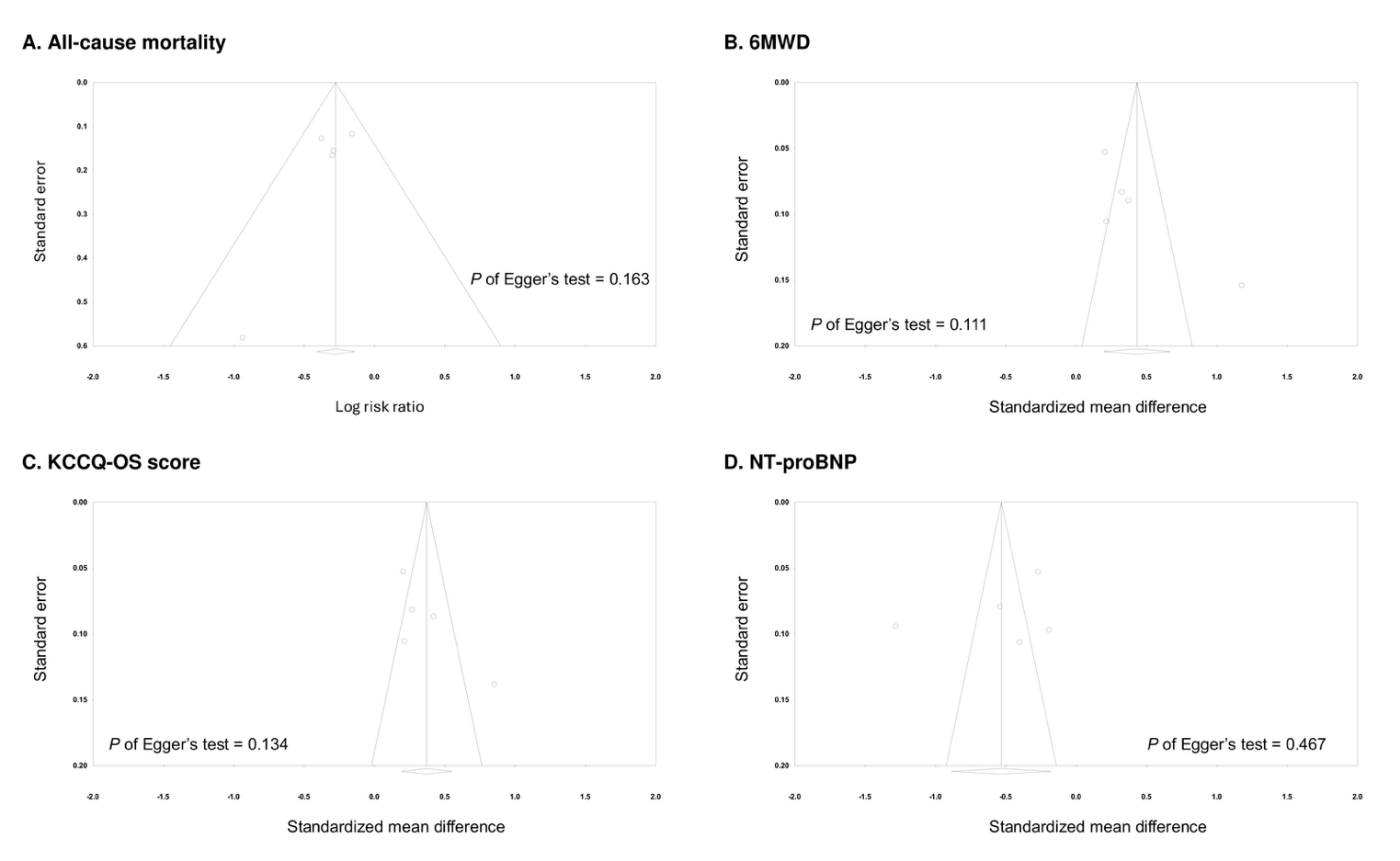
